# Galectin-3 is Associated with Heart Failure Incidence: A Meta-Analysis

**DOI:** 10.1101/2022.10.11.22280954

**Authors:** Basil M. Baccouche, Mattia A. Mahmoud, Corrine Nief, Karan Patel, B. Natterson-Horowitz

## Abstract

**Introduction:** Heart failure (HF) is a leading cause of death worldwide. The global prevalence of heart failure is projected to increase rapidly in the coming decades, and significant attention has turned to improving biomarker-based risk prediction of incident HF. The aim of this paper was to qualitatively and quantitatively evaluate the evidence associating levels of galectin-3 with risk of incident HF.

**Methods:** A review of PUBMED-indexed peer-reviewed literature was performed. Nine studies met inclusion criteria, and all nine had data eligible for conversion and pooling. A random-effects meta-analysis was performed using hazard ratios and 95% confidence intervals from a minimally adjusted model, a further adjusted model, and from subgroups within the further-adjusted model.

**Results:** The minimally-adjusted model provided a HR of 1.97 (95% CI 1.74-2.23) when comparing the top quartile of log-gal-3 to the bottom quartile. The further-adjusted model provided a HR of 1.32 (95% CI 1.21-1.44) for the same comparison. The positive, significant association was conserved during sensitivity analysis.

**Conclusion:** There is a significant positive association between levels of circulating galectin-3 and risk of incident heart failure. Given the complex mechanistic relationship between galectin-3 and cardiovascular pathophysiology, further investigation is recommended for possible implementation of galectin-3 into clinical risk prediction models.

## INTRODUCTION

### The Burden of Heart Failure

Cardiovascular disease (CVD) is the most common cause of death in the United States^1,2^. Heart failure (HF) is a chronic progressive form of CVD wherein ventricular filling or ejection of blood is impaired^3–6^. Heart failure is a pandemic affecting tens of millions, with high morbidity and mortality^7^. The global prevalence of heart failure is projected to increase rapidly in the coming decades^7^.

### Galectin-3: Protein and Biomarker

Galectin-3, a protein of the galectin family causally responsible for several physiological (and pathophysiological) processes within the cardiovascular system relating to fibrosis, atherosclerosis, and heart failure, has emerged as a potential biomarker for incidence of certain cardiovascular diseases^8,9^. Galectin-3 is expressed both intracellularly and extracellularly, and thus circulating levels of galectin-3 can be measured from serum or plasma samples^9^.

As many developing nations experience a shift towards a higher burden of non-communicable, chronic diseases, the importance of improving biomarker-based heart failure risk prediction continues to grow^10^. Although successful targeted pharmacological therapeutics have been developed for certain subtypes of HF, few effective therapies exist for other subtypes, notable among them HFpEF^11^. A potential limiting factor for the development of both preventative and acute therapies may be incomplete biomarker-based understanding of this relationship.

The purpose of this article is to rigorously evaluate and quantify the published evidence associating galectin-3 levels with incidence of heart failure. This is done to meet two primary objectives. The first objective is to reduce uncertainty surrounding the role of galectin-3 in incident heart failure via meta-analysis of the growing amount of evidence associating galectin-3 and incident heart failure. As a result of establishing an association between galectin-3 levels and incident heart failure, the second objective is to support future investigation of galectin-3 as a possible predictive biomarker of incident heart failure, a clinically useful possibility which has thus far been met with limited inquiry. Studies have been performed with mixed results assessing the value of galectin-3 in predicting mortality in patients with HF^8^. To date, the authors have identified no review and meta-analysis of the literature associating galectin-3 levels and incident HF in the peer-reviewed literature.

## METHODS

### Search Strategy

A review examining the evidence associating galectin-3 and incidence of heart failure was performed using the Sciome Workbench for Interactive computer-Facilitated Text-mining (SWIFT)-Review, which uses statistical text mining to sort search results for high-efficiency manual screening^12,13^. The following search terms were processed using the United States National Library of Medicine’s PubMed database: (HF Incidence OR heart failure incidence) AND (galectin-3 OR gal-3). No restrictions were applied to the search.

The 171 results were manually screened by title and abstract using our predefined inclusion criteria (Table 1) and in accordance with the reproducible PRISMA-compatible flow process (Figure 1).

**Table 1:** Study inclusion criteria.

| **Inclusion Criteria** |
| --- |
| -Consists of primary research |
| -Includes the exposure galectin-3 |
| -Includes the outcome incident HF |
| -Reports a measure of association (OR, HR, RR) |
| -HF clearly defined and diagnosed |
| -Community-based population |
| -Conducted on human subjects |
| -English language |
| -Full-text freely available to University of Cambridge |

**Figure 1:**
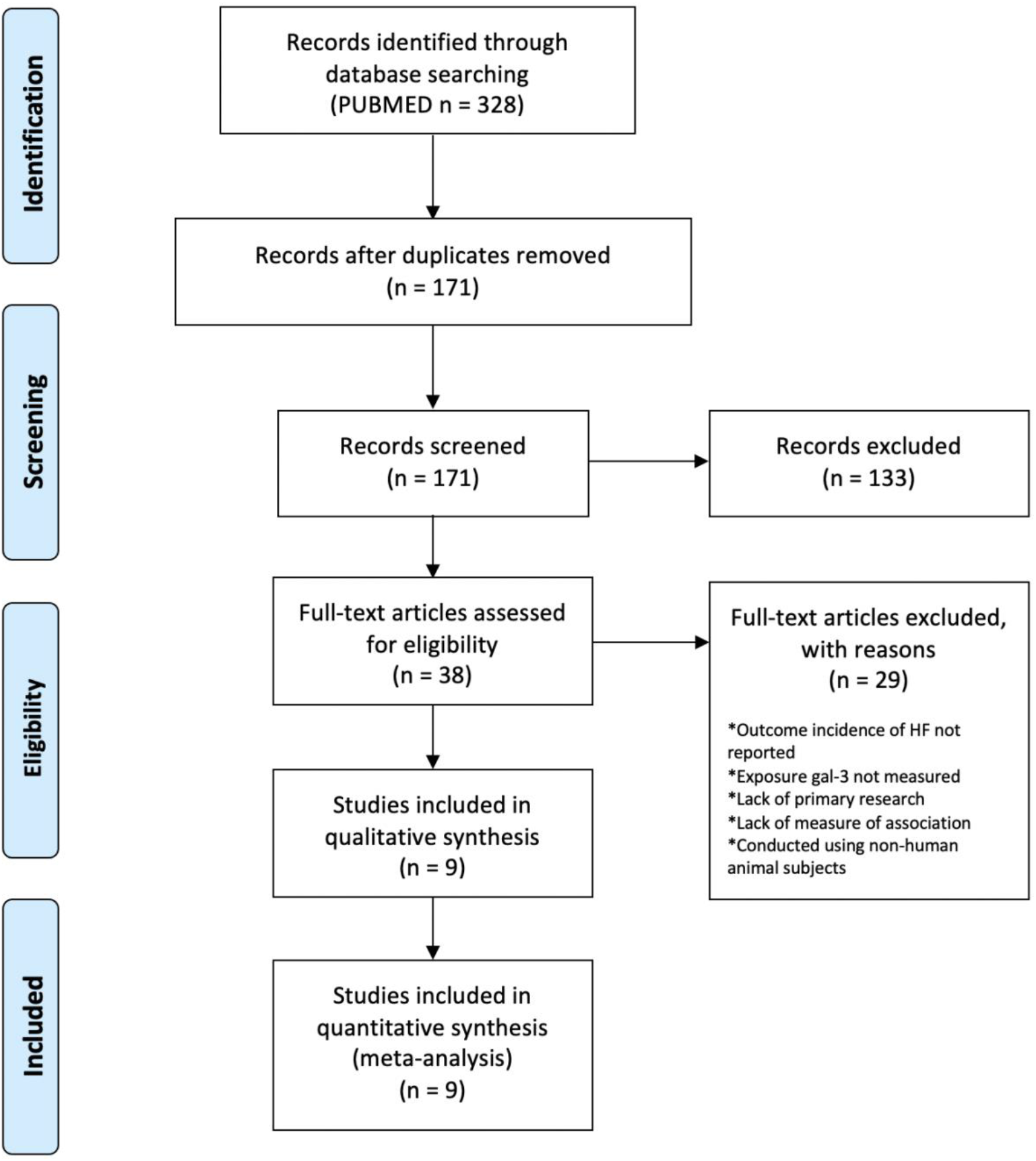
Reproducible, PRISMA-compatible review workflow^14^.

If multiple papers used the same population cohort, the study with the most relevant data was included. The study by de Boer et al. in 2018 (Table 2) includes minor overlap from community-based populations used for other studies (FHS and PREVEND)^15^. However, descriptive data for novel study populations CHS and MESA were less comprehensive and therefore less suitable for meta-analysis compared to the pooled overall data. Supplementary materials were consulted when performing full-text evaluation.

**Table 2:**
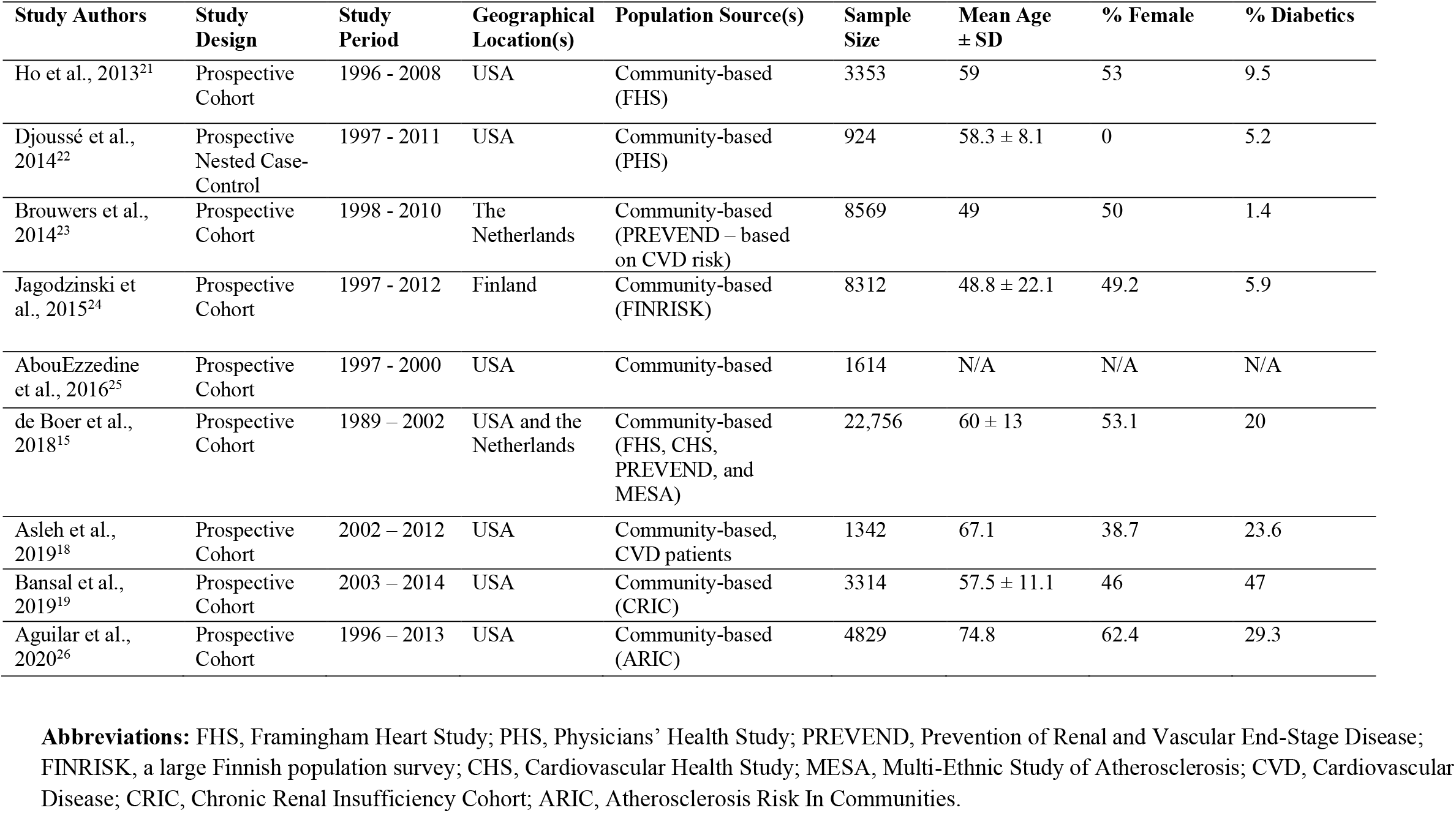
Characteristics of included studies.

### Data Collection and Quality Assessment

Study design characteristics and relevant data were manually extracted from full-text review (including supplementary materials) of included studies. Galectin-3 data were collected from the results of reported serum or plasma galectin-3 assays. Incidence of heart failure was considered the outcome of interest, defined by the included studies using respective study criteria or by hospitalization due to HF. Samples were drawn from general populations and from populations with pre-existing kidney disease, diabetes, or myocardial infarction. Hazard ratios and odds ratios were considered valid measures of association between galectin-3 and HF incidence. For rare disease, odds ratios and hazard ratios can be used interchangeably.

The Newcastle-Ottawa Scale was used to evaluate the quality of included non-randomized studies (Appendix IV). Low, moderate, and high quality studies were given scores between 0-3, 4-6, and 7-9, respectively. The complete results of this evaluation are shown in Appendix V.

### Statistical Analysis

Statistical analysis of study results was done using a random effects meta-analysis due to inter-study heterogeneity. 1 standard-deviation log-gal-3-based hazard ratios and tertile-based hazard ratios were converted to quartile-based hazard ratios, on the assumption that the log-risk ratio is linear and that log-gal-3 is normally distributed^16^.

Heterogeneous hazard ratios across studies were converted and standardized for the purpose of meta-analysis using the *riskconv* command in STATA 16.1^17^. All meta-analyses were performed using STATA 16.1. Two-sided p-values below 0.05 were considered significant unless otherwise indicated. A risk ratio whose lower 95% confidence interval remains above a value of 1 is also considered significant.

Low numbers of studies with appropriate subgroups prevented meaningful stratified meta-analysis by sex, pre-existing CVD, or location. Sensitivity analyses were performed by location, pre-existing cardiovascular disease, and to exclude outliers

## RESULTS

### Overview of Included Studies

Nine studies were included at the conclusion of the literature review, consisting of eight prospective cohort studies and 1 prospective nested case-control study. Sample sizes ranged from 924 to 22,756 participants, and the mean participant age ranged from 48.8 to 74.8 years. Most studies included diabetics in their patient population, but only one study^18^ drew from a patient population with pre-existing CVD. Another study^19^ drew from a patient population with chronic renal insufficiency. Studies were conducted in the United States and Europe. Study characteristics are displayed in Table 2.

All studies measured incident HF as an outcome of interest, and the number of HF events ranged from 166 to 2095, as shown in Table 3. In eight of the nine studies, heart failure was clearly defined using the Framingham criteria, ESC guidelines, MORGAM/AHA criteria, or ICD-9 codes (Table 3). Only one study^19^ used an unspecified “standardized clinical criteria’’ to define HF. Seven studies measured galectin-3 using an enzyme-linked immunosorbent assay (ELISA) manufactured by either BG Medicine or R&D Systems, and two used a chemiluminescent immunoassay (CMIA) manufactured by Abbott Diagnostics (Table 4).

**Table 3:** Characteristics of study outcomes.

| Study Authors | Defining Outcome of Interest (Incident HF) | Number of Events | Follow-Up Length in Years | Outcome Collection | Outcome Ascertainment* |
| --- | --- | --- | --- | --- | --- |
| Ho et al., 2013 | HF as confirmed by a panel of 3 physicians systematically reviewing records | 166 | 8.1 (Mean) | Outpatient and hospitalization records | Framingham criteria |
| Djoussé et al., 2014 | Self-reporting of HF by patients (who are themselves physicians) | 462 | 20+ (Maximum) | Annual follow-up questionnaires and (limited) chart review | Framingham criteria |
| Brouwers et al., 2014 | HF as identified by researchers using ESC guidelines | 374 | 12.5 (Median) | Outpatient and hospitalization records | ESC guidelines |
| Jagodzinski et al., 2015 | HF as identified by researchers using MORGAM/AHA criteria | 641 | 15 (Maximum) | Finnish national health care registry | MORGAM and AHA criteria |
| AbouEzzedine et al., 2016 | HF as defined by direct physician diagnosis or by trained nurse abstractors from medical records using Framingham criteria/ICD-9 codes | 173 | 11 (Median) | Rochester Epidemiology Project medical records | Framingham criteria, ICD-9 code 402 or 428 |
| de Boer et al., 2018 | FHS: HF as confirmed by a panel of 3 physicians systematically reviewing records<br>CHS: Physician diagnosis of patients during follow-up<br>META: HF as indicated by death certificates, hospitalization records, or interviews<br>PREVEND: HF as identified by researchers using ESC guidelines | 2095 | 12 (Mean) | FHS: Outpatient and hospitalization records<br>CHS: Extensive physical and laboratory examinations and follow-up<br>MESA: Death certificates, hospitalization records, interviews<br>PREVEND: Outpatient and hospitalization records | FHS: Framingham criteria<br>CHS: CHS criteria, ICD-9 codes 427-429.9<br>MESA: Identified from records<br>PREVEND: ESC guidelines |
| Asleh et al., 2019 | HF as identified by researchers from records using Framingham criteria/ICD-9 codes | 368 | 5.4 (Mean) | Rochester Epidemiology Project medical records | Framingham criteria, ICD-9 code 428 |
| Bansal et al., 2019 | 2 researchers agreed upon a “probable” or “definite” HF diagnosis from follow-up questionnaires and medical records | 477 | 7.9 (Median) | Hospitalization records and biannual questionnaire | “accepted standardized clinical criteria” (unspecified) |
| Aguilar et al., 2020 | HF hospitalizations adjudicated by experts, and trained personnel abstracted medical/death records for HF | 1599 | 18 (Median) | Hospitalization records and annual participant reporting | ICD-9 code 428 |

\*For Framingham criteria, see Appendix I. For ESC guidelines, see Appendix II. For CHS criteria, see Appendix III. The ICD-9 (International Classification of Diseases, Ninth Revision) is an internationally adopted classification scheme used to collect, process, classify, and present worldwide mortality statistics<sup>27</sup>. Diagnosis of HF derived from the MORGAM and AHA criteria, in conjunction with Finnish national health records, have been demonstrated to be of high specificity and validity<sup>28</sup>.
**Abbreviations:** HF, Heart Failure; ESC, European Society of Cardiology; AHA, American Heart Association; ICD-9, International Classification of Diseases, Ninth Edition; FHS, Framingham Heart Study; CHS, Cardiovascular Health Study; MESA, Multi-Ethnic Study of Atherosclerosis; PREVEND, Prevention of Renal and Vascular End-Stage Disease.

**Table 4:** Characteristics of study assays.

| **Study Authors** | **Sample Type** | **Storage Duration (Years)** | **Storage Temperature (°C)** | **Galectin Measurement Method\*** | **Assay Manufacturer** |
| --- | --- | --- | --- | --- | --- |
| Ho et al., 2013 | Plasma | 14-16 | -70 | ELISA | BG Medicine, Waltham, MA |
| Djoussé et al., 2014 | Plasma | 20 | N/A | ELISA | R&D Systems, Minneapolis, MN |
| Brouwers et al., 2014 | Plasma | 16 | -80 | ELISA | BG Medicine, Waltham, MA |
| Jagodzinski et al., 2015 | N/A | 20 | -70 | CMIA | Abbot Diagnostics, Abbott Park, IL |
| AbouEzzedine et al., 2016 | N/A | N/A | N/A | ELISA | BG Medicine, Waltham, MA |
| de Boer et al., 2018 | N/A | N/A | N/A | ELISA | BG Medicine, Waltham, MA |
| Asleh et al., 2019 | Plasma | 7-17 | -70 | ELISA | BG Medicine, Waltham, MA |
| Bansal et al., 2019 | Plasma | 6-11 | -70 | ELISA | R&D Systems, Minneapolis, MN |
| Aguilar et al., 2020 | Plasma | 4-20 | -70 | CMIA | Abbot Diagnostics, Abbott Park, IL |

\*An enzyme-linked immunosorbent assay (ELISA) is a highly sensitive test used to detect and quantify substances including antibodies, antigens, proteins, and hormones in biological samples<sup>29</sup>. A chemiluminescent immunoassay (CMIA) is another type of immunoassay used for detecting substances in biological samples wherein the label (the indicator of the reaction) is a luminescent molecule<sup>30</sup>.

Main findings were reported as hazard ratios in the eight prospective cohort studies and as an odds ratio in the prospective nested case-control study (Table 5). Studies in which the hazard ratio was reported by comparing 1 standard deviation log-gal-3 increase, the 3rd tertile of gal-3 compared to bottom tertile, or linear gal-3 doubling to incident HF were converted to 4th quartile-to-bottom-quartile log-gal-3 to incident HF hazard ratios for the purpose of meta-analysis^20^. The converted values are displayed in Appendix VI.

**Table 5:** Main findings of included studies. *Converted values can be found in Appendix VI.

| **Study Authors** | **Minimum Adjusted Model HR [95% CI] (Covariates)** | **Further Adjusted Model HR [95% CI]** | **Comparison\*** | **Covariates in Further Adjusted Model** |
| --- | --- | --- | --- | --- |
| Ho et al., 2013 | 1.39<br>[1.17-1.65]<br>(Age, sex) | 1.23<br>[1.04-1.47] | 1 SD log-Gal-3 increase and incident HF | Age, sex, SBP, anti-HTN treatment, BMI, diabetes mellitus, smoking, CAD, AF, BNP, and valvular heart disease. |
| Djoussé et al., 2014 | **Odds Ratio:**<br>1.60<br>[1.07-2.40]<br>(AF, BMI, diabetes) | **Odds Ratio:**<br>1.57<br>[1.03-2.39] | 4<sup>th</sup> quartile of log-gal-3 compared to bottom quartile and incident HF | AF, BMI, diabetes, HTN, log(hsCRP), alcohol, exercise, and smoking. |
| Brouwers et al., 2014 | 1.18<br>[1.03-1.36]<br>(Age, sex) | 1.10<br>[0.95-1.28] | Linear gal-3 doubling and incident HF | Age, sex, BMI, smoking, SBP, plasma glucose, total cholesterol. |
| Jagodzinski et al., 2015 | 1.46<br>[1.13-1.90]<br>(Region of Finland) | 1.22<br>[0.94-1.58] | 4<sup>th</sup> quartile of gal-3 compared to bottom quartile and incident HF | Region of Finland, HDL/total cholesterol, SBP, anti-HTN treatment, smoking, diabetes, valvular heart disease, and eGFR. |
| AbouEzzedine et al., 2016 | 1.50<br>[1.32-1.71]<br>(Unadjusted) | 1.17<br>[0.99-1.39] | 1 SD log-Gal-3 increase and incident HF | Age, sex, BMI, eGFR, HTN, SBP, CAD, and diabetes mellitus. |
| de Boer et al., 2018 | N/A | 1.07<br>[1.02-1.12] | 1 SD log-Gal-3 increase and incident HF | Pooled and adjusted for age, sex, race/ethnicity, SBP, HTN, BMI, diabetes, smoking, LVH, left bundle branch block, and previous MI.<br>(CHS: Adjusted for age, sex, race/ethnicity, previous MI, BMI, anti-HTN treatment, SBP, smoking, LVH, left bundle branch block, and diabetes). |
| Asleh et al., 2019 | 3.46<br>[2.51-4.78]<br>(Age, sex) | 2.25<br>[1.61-3.15] | 3<sup>rd</sup> tertile of gal-3 compared to bottom tertile and incident HF | Age, sex, Charlson comorbidity index, maximum troponin T, and history of HF. |
| Bansal et al., 2019 | 1.17<br>[1.05-1.30]<br>(Age, sex, race, diabetes mellitus, CVD, smoking, 24-hr urinary protein, eGFR, SBP, BMI, LDL cholesterol, HDL cholesterol) | 1.12<br>[1.00-1.24] | 1 SD log-Gal-3 increase and incident HF | Age, sex, race, diabetes mellitus, CVD, smoking, 24-hr urinary protein, eGFR, SBP, BMI, LDL cholesterol, HDL cholesterol, ACE inhibitors/blockers, diuretics, beta-blockers, phosphate, parathyroid hormone, FGF-23. |
| Aguilar et al., 2020 | 3.90<br>[2.47-6.17]<br>(Age, sex, race, SBP, anti-HTN treatment, smoking, diabetes mellitus, BMI, and heart rate) | 1.93<br>[1.15-3.24] | 1 SD log-Gal-3 increase and incident HF | Age, sex, race, SBP, anti-HTN treatment, smoking, diabetes mellitus, BMI, heart rate, eGFR, log NT-proBNP, and log high sensitivity cardiac troponin T. |

**Abbreviations:** HR, hazard ratio; SD, standard deviation; HF, heart failure; SBP, systolic blood pressure; HTN, hypertension; BMI, body-mass index; CAD; coronary artery disease; AF, atrial fibrillation; BNP, brain natriuretic peptide; CRP, C-reactive protein; HDL, high-density lipoprotein; eGFR, estimated glomerular filtration rate; LVH, left ventricular hypertrophy; MI, myocardial infarction; LDL, low-density lipoprotein; ACE, angiotensin-converting enzyme; FGF, fibroblast growth factor; NT-proBNP, N-terminal pro B-type natriuretic peptide.

The Newcastle-Ottawa Scale was used to quantify the methodological quality of each of the studies included (Appendix IV & V). All included studies received a score of 7 or higher, indicating high quality. Limitations of each individual study are listed in Discussion (Table 6).

**Table 6:**
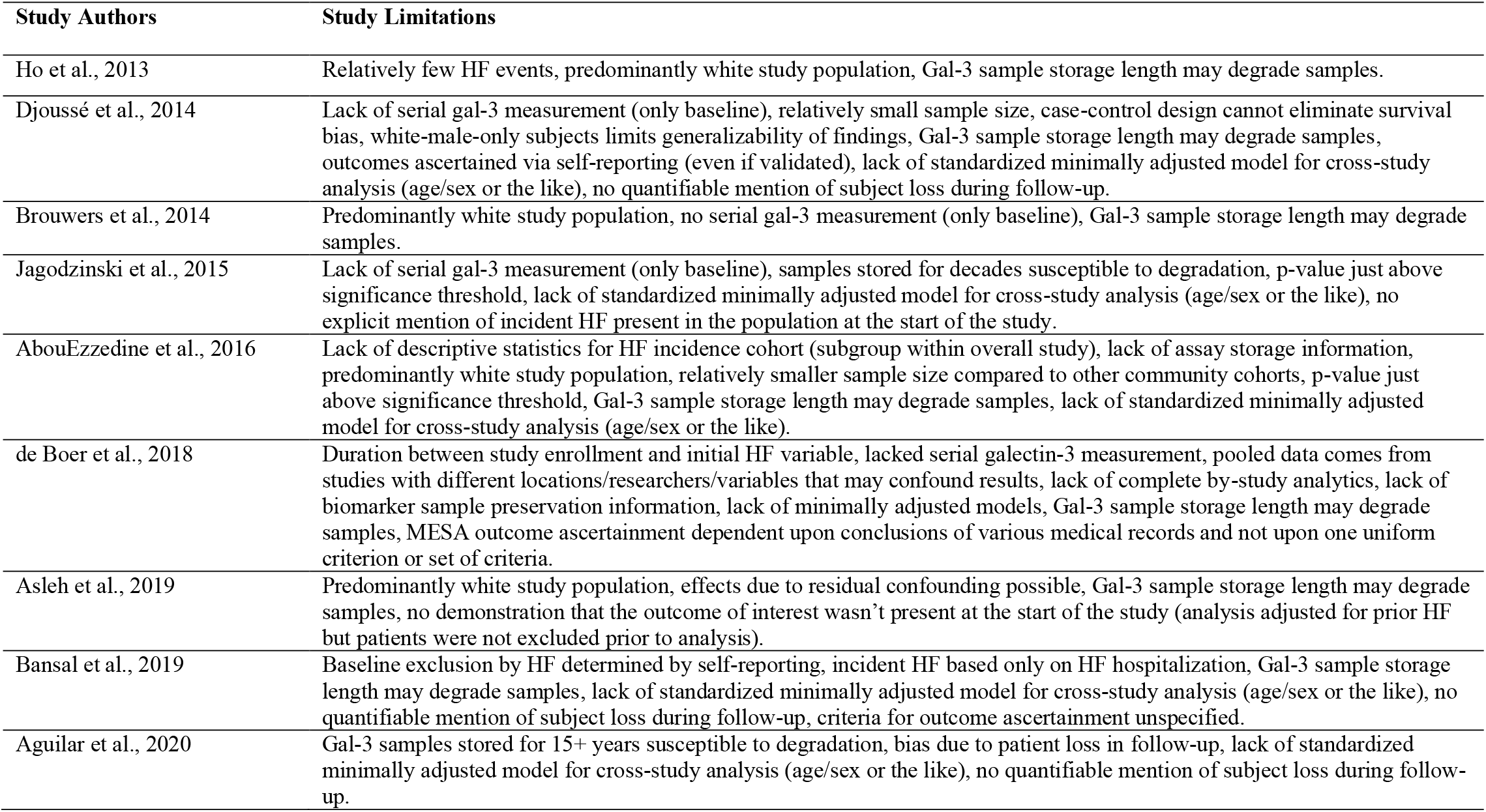
Limitations of Included Studies.

| **Study Authors** | **Study Limitations** |
| --- | --- |
| Ho et al., 2013 | Relatively few HF events, predominantly white study population, Gal-3 sample storage length may degrade samples. |
| Djoussé et al., 2014 | Lack of serial gal-3 measurement (only baseline), relatively small sample size, case-control design cannot eliminate survival bias, white-male-only subjects limits generalizability of findings, Gal-3 sample storage length may degrade samples, outcomes ascertained via self-reporting (even if validated), lack of standardized minimally adjusted model for cross-study analysis (age/sex or the like), no quantifiable mention of subject loss during follow-up. |
| Brouwers et al., 2014 | Predominantly white study population, no serial gal-3 measurement (only baseline), Gal-3 sample storage length may degrade samples. |
| Jagodzinski et al., 2015 | Lack of serial gal-3 measurement (only baseline), samples stored for decades susceptible to degradation, p-value just above significance threshold, lack of standardized minimally adjusted model for cross-study analysis (age/sex or the like), no explicit mention of incident HF present in the population at the start of the study. |
| AbouEzzedine et al., 2016 | Lack of descriptive statistics for HF incidence cohort (subgroup within overall study), lack of assay storage information, predominantly white study population, relatively smaller sample size compared to other community cohorts, p-value just above significance threshold, Gal-3 sample storage length may degrade samples, lack of standardized minimally adjusted model for cross-study analysis (age/sex or the like). |
| de Boer et al., 2018 | Duration between study enrollment and initial HF variable, lacked serial galectin-3 measurement, pooled data comes from studies with different locations/researchers/variables that may confound results, lack of complete by-study analytics, lack of biomarker sample preservation information, lack of minimally adjusted models, Gal-3 sample storage length may degrade samples, MESA outcome ascertainment dependent upon conclusions of various medical records and not upon one uniform criterion or set of criteria. |
| Asleh et al., 2019 | Predominantly white study population, effects due to residual confounding possible, Gal-3 sample storage length may degrade samples, no demonstration that the outcome of interest wasn't present at the start of the study (analysis adjusted for prior HF but patients were not excluded prior to analysis). |
| Bansal et al., 2019 | Baseline exclusion by HF determined by self-reporting, incident HF based only on HF hospitalization, Gal-3 sample storage length may degrade samples, lack of standardized minimally adjusted model for cross-study analysis (age/sex or the like), no quantifiable mention of subject loss during follow-up, criteria for outcome ascertainment unspecified. |
| Aguilar et al., 2020 | Gal-3 samples stored for 15+ years susceptible to degradation, bias due to patient loss in follow-up, lack of standardized minimally adjusted model for cross-study analysis (age/sex or the like), no quantifiable mention of subject loss during follow-up. |

### Meta-Analysis

After conversion, all nine included studies reported data conducive to further-adjusted meta-analysis, but only eight reported data conducive to minimally-adjusted meta-analysis. de Boer et al., 2018^15^ was excluded from the minimally adjusted meta-analysis because they did not publish a minimally adjusted model.

Each study individually demonstrated a positive association between 4th-quartile log-gal-3 levels and risk of heart failure. The minimally adjusted random-effects model (Figure 2) provided a hazard ratio of 1.97 (95% CI 1.74-2.23) with heterogeneity statistic I^2^ = 87.3% (95% CI 76.7-91.9), indicating high heterogeneity. The maximally adjusted random-effects model (Figure 3) provided a hazard ratio of 1.32 (95% CI 1.21-1.44) with heterogeneity statistic I^2^ = 61.6% (95% CI 0-79.7), indicating moderate heterogeneity. Both the minimally and further-adjusted meta-analyses demonstrate a robust, significant positive association.

**Figure 2:**
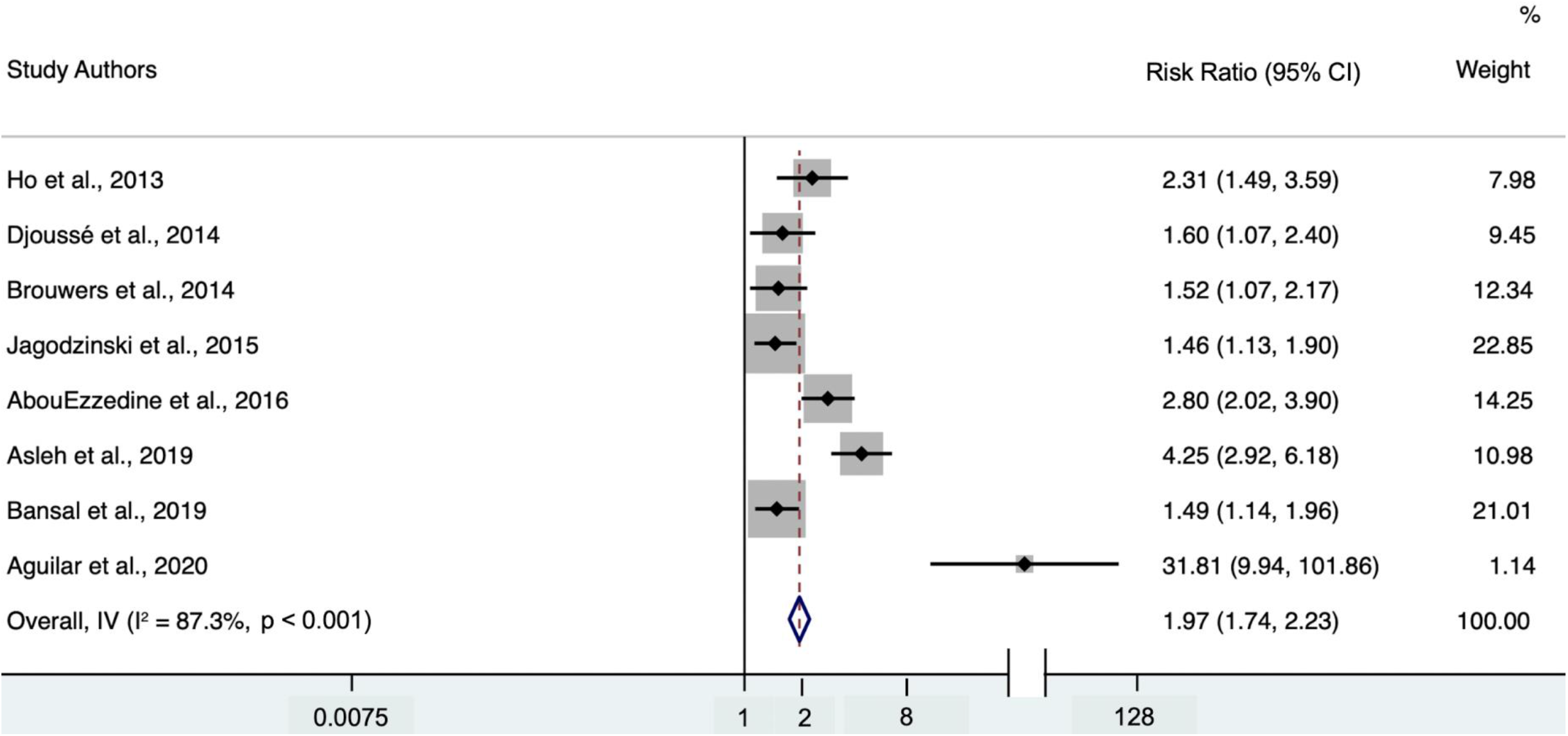
Forest plot of minimally adjusted model hazard ratios for the association between 4th-quartile-to-bottom-quartile log-gal-3 and incident HF. Study weights (represented by the grey boxes) are from random-effects analysis. 95% confidence interval for the heterogeneity statistic I^2^: (76.7-91.9).

**Figure 3:**
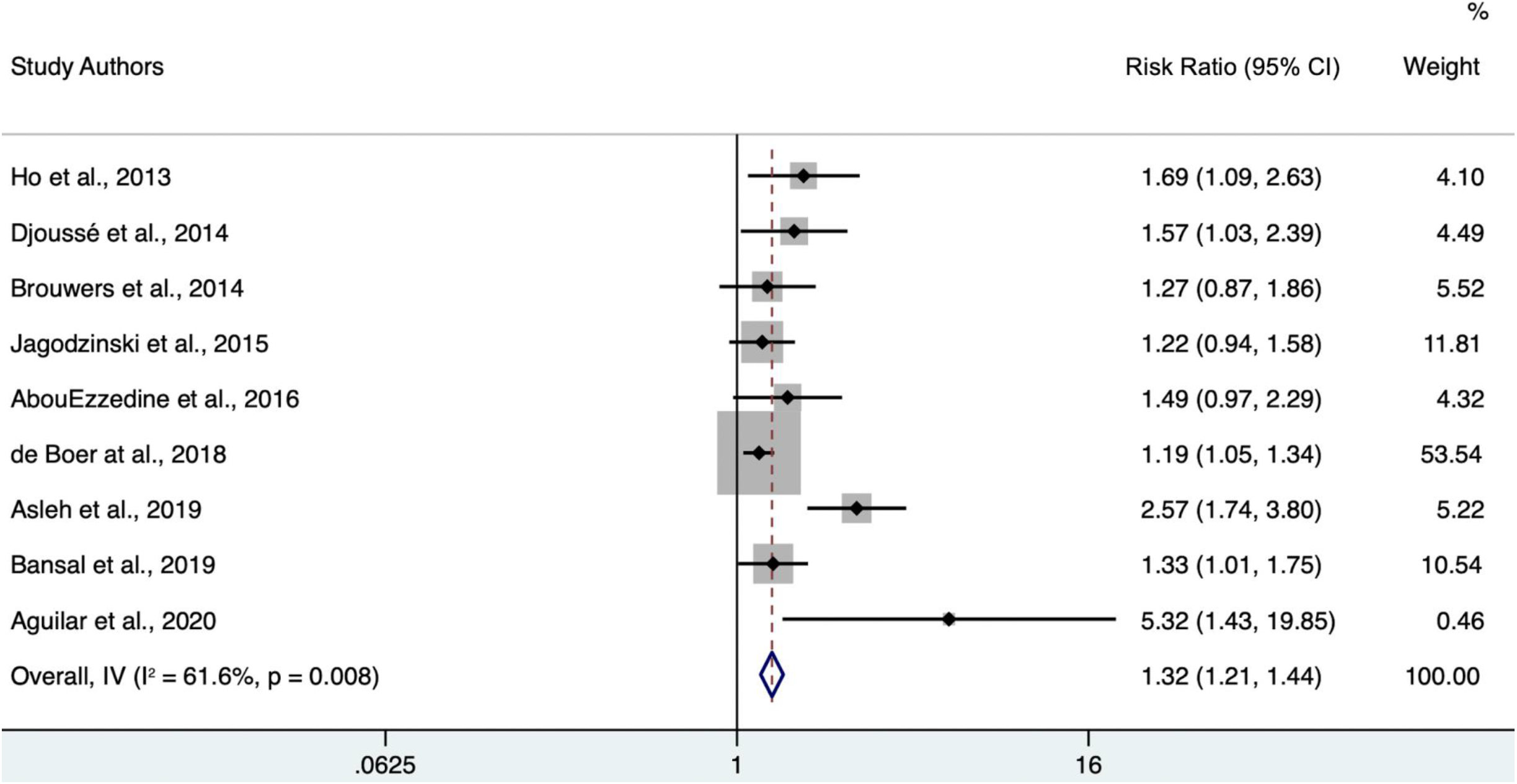
Forest plot of further adjusted model hazard ratios for the association between 4th-quartile-to-bottom-quartile log-gal-3 and incident HF. Study weights (represented by the grey boxes) are from random-effects analysis. 95% confidence interval for the heterogeneity statistic I^2^: (0-79.7).

The study by Aguilar et al. in 2020^26^ was an obvious outlier. Due to its low weight, the overall further-adjusted hazard ratio (1.31 (95% CI 1.20-1.43)) did not change significantly upon its exclusion, as shown in Figure 4. However, outlier exclusion did reduce the observed heterogeneity (I^2^ = 57.5% (95% CI 0-78.8)).

**Figure 4:**
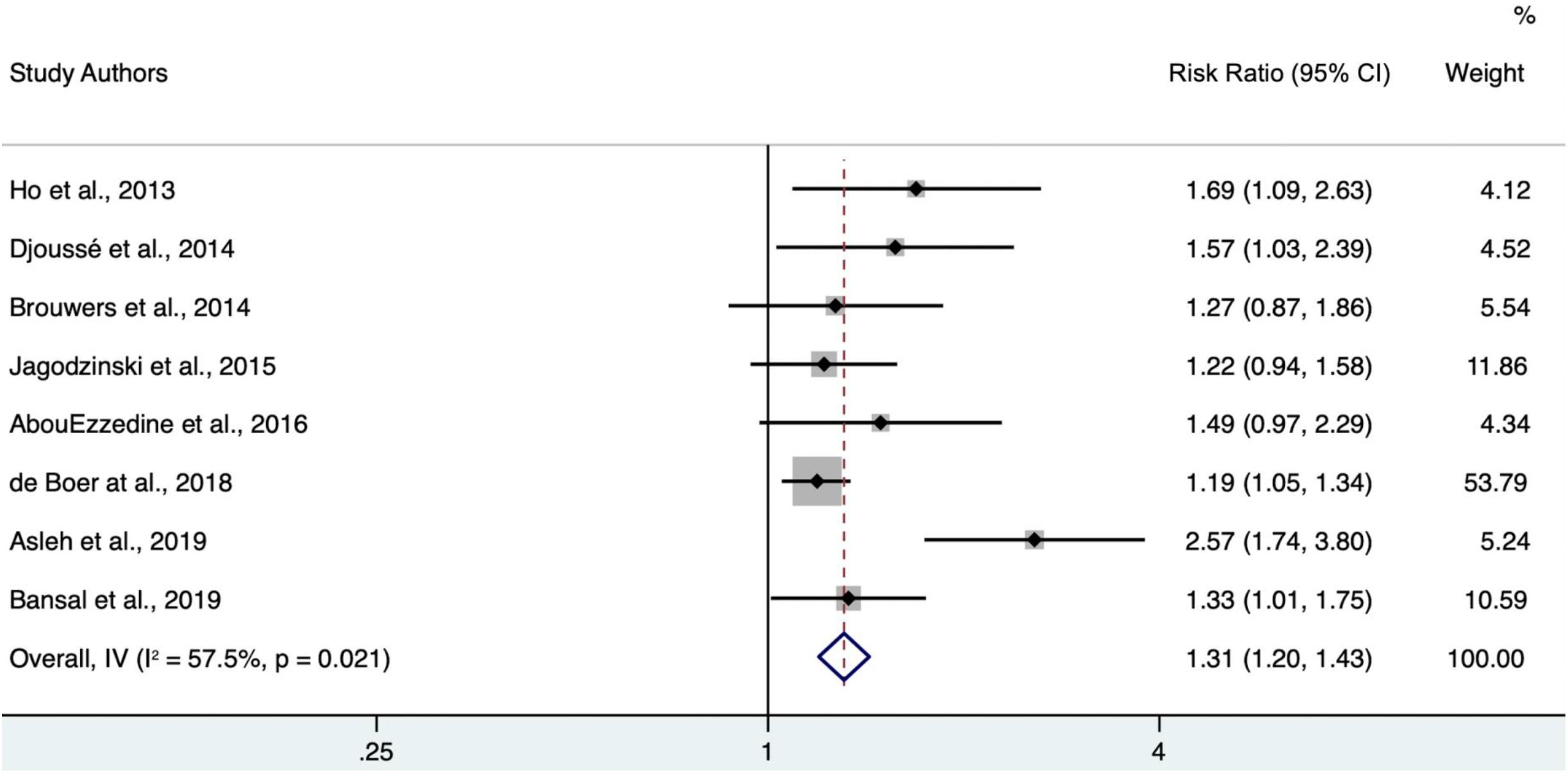
Forest plot of further adjusted model hazard ratios for the association between 4th-quartile-to-bottom-quartile log-gal-3 and incident HF, excluding the outlier Aguilar et al., 2020. Study weights (represented by the grey boxes) are from random-effects analysis. 95% confidence interval for the heterogeneity statistic I^2^: (0-78.8).

### Sensitivity Meta-Analysis

All sensitivity meta-analyses were performed using further-adjusted models. Only one study^18^ utilized a study population with pre-existing CVD. Upon exclusion of this study (Figure 5), the overall hazard ratio was very modestly lowered from 1.32 (95% CI 1.21-1.44) to 1.27 (95% CI 1.16-1.39). Heterogeneity statistic I^2^ = 22% (95% CI 0-65.3), indicating low heterogeneity.

**Figure 5:**
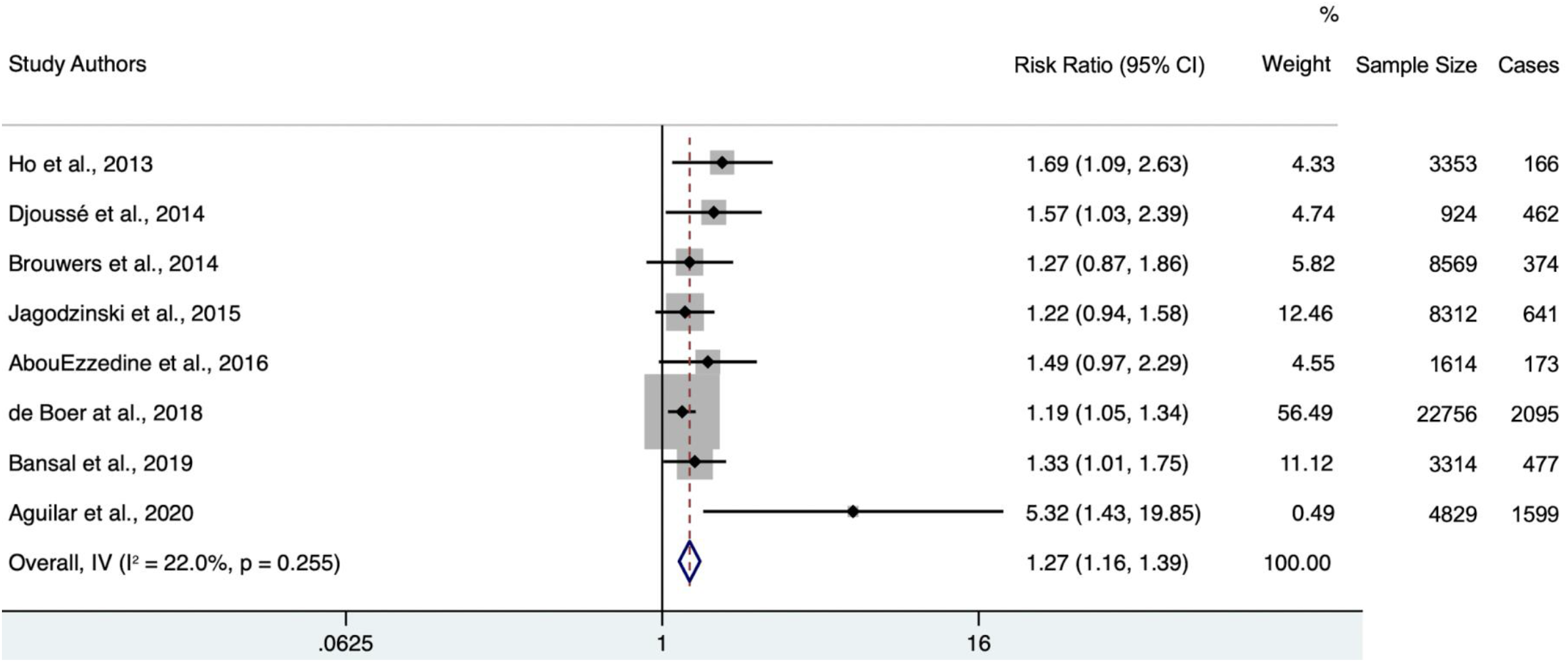
Forest plot of further adjusted model hazard ratios for the association between 4th-quartile-to-bottom-quartile log-gal-3 and incident HF, excluding study populations with pre-existing CVD. Study weights (represented by the grey boxes) are from random-effects analysis. 95% confidence interval for the heterogeneity statistic I^2^: (0-65.3).

Six of the nine included studies utilized study populations exclusively within the United States. Meta-analysis of USA-based studies (Figure 6) notably elevated the overall hazard ratio from 1.32 (95% CI 1.21-1.44,) to 1.65 (95% CI 1.40-1.65). Heterogeneity statistic I^2^ = 53% (95% CI 0-79.3), indicating moderate heterogeneity.

**Figure 6:**
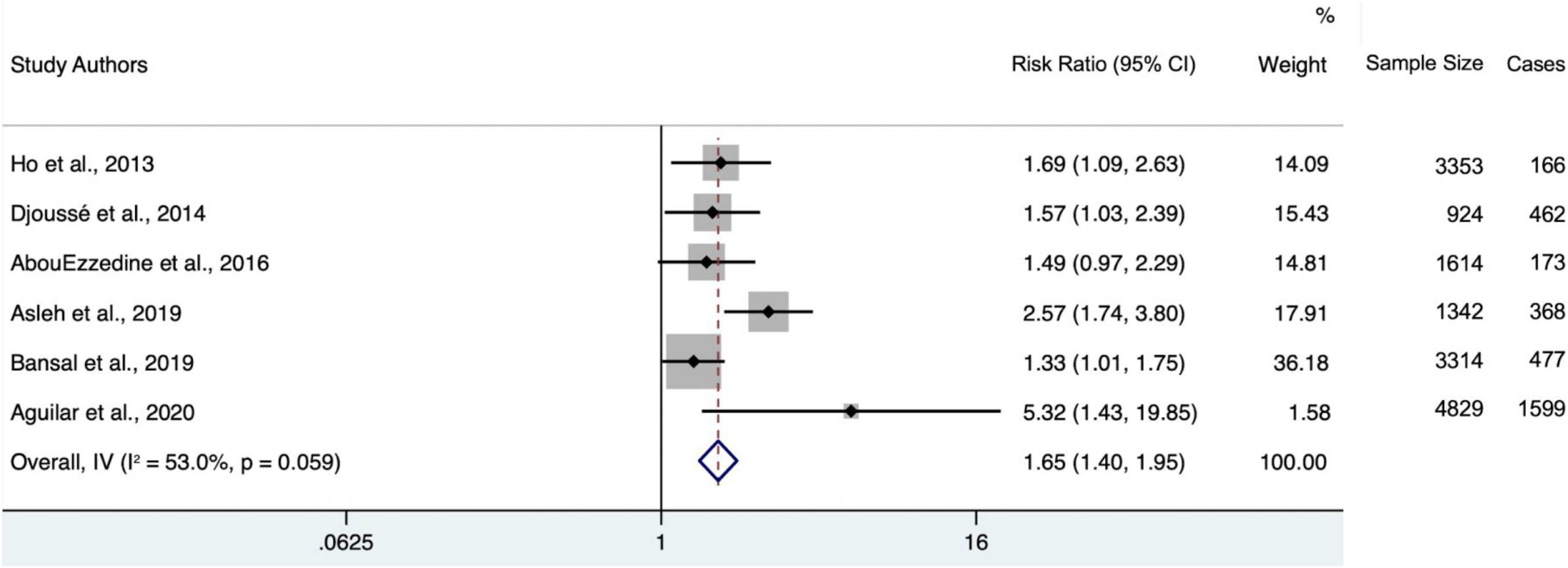
Forest plot of further adjusted model hazard ratios for the association between 4th-quartile-to-bottom-quartile log-gal-3 and incident HF, including only studies whose patient populations are exclusively within the United States. Study weights (represented by the grey boxes) are from random-effects analysis. 95% confidence interval for the heterogeneity statistic I^2^: (0-79.3).

Of all included studies, de Boer et al., 2018^15^ had the largest weight percentage within the further-adjusted meta-analysis (53.54%). Exclusion of this study (Figure 7) elevated the overall hazard ratio from 1.32 (95% CI 1.21-1.44) to 1.48 (95% CI 1.30-1.69). Heterogeneity statistic I^2^ = 53.5% (95% CI 0-77.2), indicating moderate heterogeneity.

**Figure 7:**
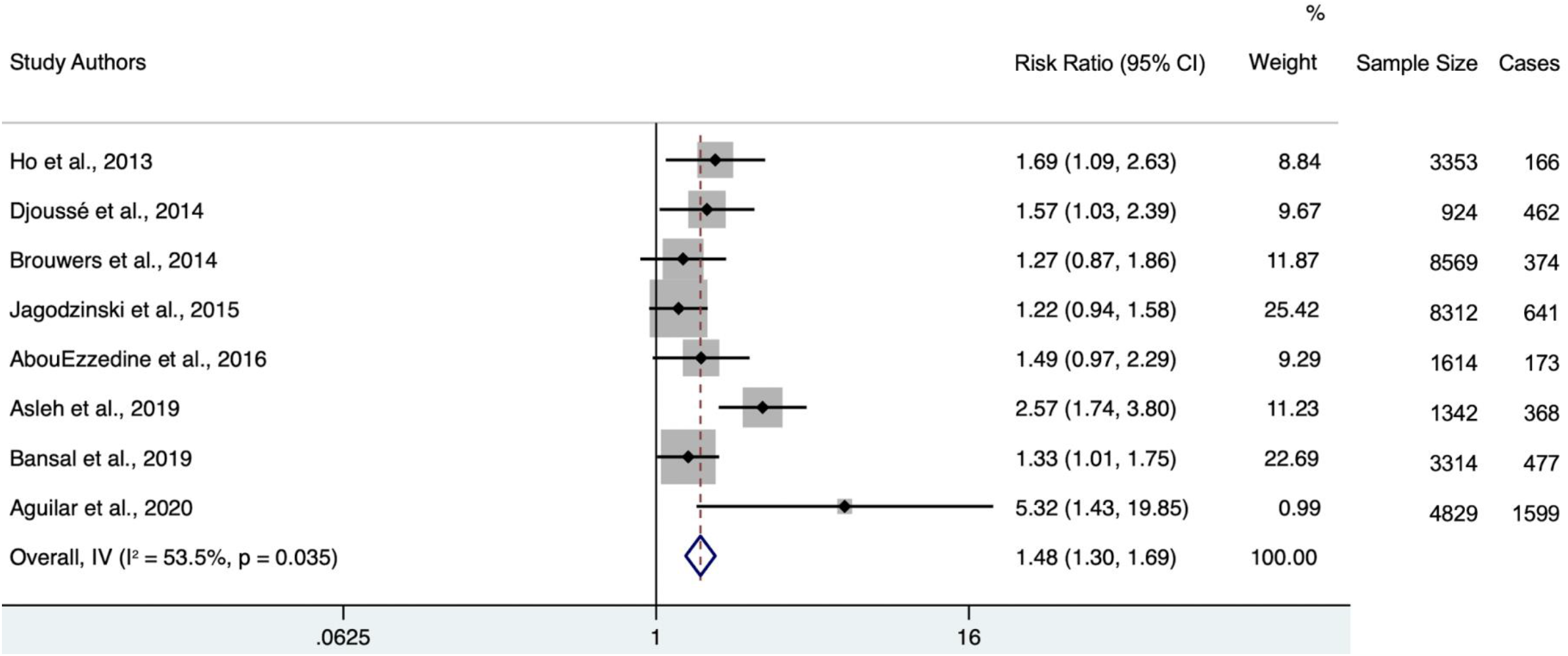
Forest plot of further adjusted model hazard ratios for the association between 4th-quartile-to-bottom-quartile log-gal-3 and incident HF, excluding the study with the highest weight (de Boer at al., 2018). Study weights (represented by the grey boxes) are from random-effects analysis. 95% confidence interval for the heterogeneity statistic I^2^: (0-77.2).

## DISCUSSION

### Summary of Findings

This review and meta-analysis demonstrates a significant association between the top-to bottom-quartile log-galectin-3 and risk of heart failure. Adjusted meta-analysis presents a 32% higher HF risk in individuals with top-quartile log-galectin-3 levels compared to those in the bottom quartile. Sensitivity analysis revealed that the significant positive association was conserved or even elevated upon outlier removal, location standardization, and removal of the study population with pre-existing CVD. Heterogeneity between studies was notable, but decreased upon adjustment and decreased further upon outlier removal and pre-existing CVD removal.

### Study Limitations

The authors did not review preprint servers or non-PUBMED-indexed journals. As a result, it is possible that relevant data (even if not yet peer-reviewed) was overlooked.

The study by de Boer et al. in 2018^15^ pooled data from four prospective cohort studies, two of which (FHS and PREVEND) had participants included in other studies. However, data on the individual unique cohorts (CHS and MESA) had significant descriptive omissions compared to information published on the pooled results, and so pooled data were included despite minor overlap.

Across the nine studies, different sets of criteria were applied to diagnose HF, introducing possible inter-study heterogeneity in HF diagnosis. Within the MESA cohort, which contributed to the pooled de Boer et al.^15^ data, outcome ascertainment was dependent upon the conclusions of various medical records and not standardized to one uniform set of criteria, introducing possible inter-record heterogeneity in HF diagnosis.

The studies included in the minimally adjusted meta-analysis are heterogeneous in their choice of covariate adjustment; while a plurality adjusted for age/sex, others were unadjusted or made significant adjustments. The studies included in the further-adjusted meta-analyses are also heterogeneous in their choice of covariate adjustment.

Heterogeneity between studies as defined by the I^2^ statistic was moderate to high in most studies. Unfortunately, given the recency with which the association between galectin-3 and incident HF has been investigated, the number of studies to choose from are limited and heterogeneous.

The relative paucity of published research on the association in question meant that the number of meta-analyzed studies (9) was low. In addition, the scarcity of available research meant that our outcome of interest (incident HF) could not be further specified by subtype (HFrEF and HFpEF).

Although robust associations were observed between circulating galectin-3 levels and incident heart failure, statistical causality was not established by this study and is recommended as a promising future direction of research to resolve the ongoing debate surrounding the possible role of gal-3 in heart failure onset.

Finally, each included study had its own limitations, represented below in Table 6.

### Study Strengths

This review synthesizes and meta-analyzes data on the association between galectin-3 and incident HF. Interest in the relationship between galectin-3 exposure and incident HF outcome has only occurred within the last decade or so. As a result, this review provides an up-to-date synthesis of established knowledge. Where specified, Galectin-3 was measured ubiquitously within the same sample medium (plasma), and assay manufacturers were reputable. Individual study sample sizes are generally high. All included studies had data compatible for meta-analysis, and there were no exclusions in the further-adjusted model. In addition, further-adjusted models were rigorously subjected to sensitivity analysis by location, pre-existing cardiovascular disease, and outlier studies. The overall association was robust and highly significant. Finally, application of the Newcastle-Ottawa Scale to our studies (Appendix IV & V) demonstrated that all nine included studies were considered to be of “high” quality.

### Implications

Heart failure is a complex physiological dysfunction with multiple contributing factors and a variety of symptoms. The highly significant overall positive association between galectin-3 and incident HF lends credence to its use as a tool to predict and prevent HF onset. However, given a top-quartile-to-bottom hazard ratio of 1.32, it is unlikely that measuring levels of galectin-3 alone will provide a comprehensive predictive model. It has been demonstrated that, although the exposure galectin-3 is positively associated with the outcome incident HF, adjusting for other biomarkers such as B-type natriuretic peptide (used in diagnosing HF) reduces the strength of this association^26^. It is therefore plausible that a combination of biomarkers, if correctly identified, might comprise a robust predictive model of incident HF in populations without symptomatic CVD. The results of this meta-analysis make a case for the possible inclusion of galectin-3 in that model. Further studies specifically evaluating the interaction between several of these biomarkers in predicting incident HF are recommended. As this study did not evaluate appropriate measures of risk prediction (such as a c-index), direct extrapolation of this study’s results to clinical risk prediction is limited without further research.

The significant drop in between-study heterogeneity upon exclusion of the study with pre-existing CVD may be due to possible reverse causation. However, as there was only one study with data on this specific association in CVD patients, further research is recommended. Future studies would be encouraged to draw from populations without pre-existing cardiovascular disease and to include raw data for adjustment standardization.

## Conclusion*s*

There is a significant positive association between top-quartile log-galectin-3 levels and risk of incident heart failure in community-based populations. However, notable heterogeneity between studies due to limited relevant published research warrants additional evidence. In particular, further studies on the association between galectin-3 and incident HF in studies without pre-existing CVD, further studies adjusting for other biomarkers implicated in HF risk, and further studies evaluating measures of risk association would help optimize the utility of galectin-3 as a predictive tool in clinical settings.

## Supporting information

PRISMA Checklist

## Data Availability

All data used in the study are already available from previously published studies and have been meta-analyzed in this study (see Figure references 15, 18, 19, and 21-26 for original data which has already been published)

## DECLARATIONS

### CONFLICTS OF INTEREST

The authors report no relationships that could be construed as a conflict of interest.

### FUNDING DECLARATION

There is no funding to declare.

## APPENDIX

### Appendix I. Framingham criteria for HF diagnosis^31^

**Table 1.** Criteria of CHF. ^*^.

| MAJOR CRITERIA |  |
| --- | --- |
| Paroxysmal nocturnal dyspnea or orthopnea |  |
| Neck-vein distention |  |
| Rales |  |
| Cardiomegaly |  |
| Acute pulmonary edema |  |
| S<sub>3</sub> gallop |  |
| Increased venous pressure $>16$ cm of water | |
| Circulation time $\geq 25$ sec | |
| Hepatojugular reflux |  |
| MINOR CRITERIA |  |
| Ankle edema |  |
| Night cough |  |
| Dyspnea on exertion |  |
| Hepatomegaly |  |
| Pleural effusion |  |
| Vital capacity $\downarrow \frac{1}{3}$ from maximum | |
| Tachycardia (rate of $\geq 120/\text{min}$) | |
| MAJOR OR MINOR CRITERION |  |
| Weight loss $\geq 4.5$ kg in 5 days in response to treatment | |

\*For establishing a definite diagnosis of congestive heart failure in this study, 2 major or 1 major & 2 minor criteria had to be present concurrently.

### Appendix II ESC guidelines for HF diagnosis^32^

**Table 3.1.** Definition of heart failure with preserved (HFpEF), mid-range (HFmrEF) and reduced ejection fraction (HFrEF)

| Type of HF | HFrEF | HFmrEF | HFpEF |
| --- | --- | --- | --- |
| CRITERIA | 1 Symptoms ± Signs<sup>a</sup> | Symptoms ± Signs<sup>a</sup> | Symptoms ± Signs<sup>a</sup> |
|  | 2 LVEF <40% | LVEF 40–49% | LVEF ≥50% |
|  | 3 – | 1. Elevated levels of natriuretic peptides<sup>b</sup>;<br>2. At least one additional criterion:<br>a. relevant structural heart disease (LVH and/or LAE),<br>b. diastolic dysfunction (for details see Section 4.3.2). | 1. Elevated levels of natriuretic peptides<sup>b</sup>;<br>2. At least one additional criterion:<br>a. relevant structural heart disease (LVH and/or LAE),<br>b. diastolic dysfunction (for details see Section 4.3.2). |

BNP = B-type natriuretic peptide; HF = heart failure; HFmrEF = heart failure with mid-range ejection fraction; HFpEF = heart failure with preserved ejection fraction; HFrEF = heart failure with reduced ejection fraction; LAE = left atrial enlargement; LVEF = left ventricular ejection fraction; LVH = left ventricular hypertrophy; NT-proBNP = N-terminal pro-B type natriuretic peptide.
<sup>a</sup>Signs may not be present in the early stages of HF (especially in HFpEF) and in patients treated with diuretics.
<sup>b</sup>BNP > 35 pg/ml and/or NT-proBNP > 125 pg/mL.

**Table 4.1.** Symptoms and signs typical of heart failure.

| Symptoms | Signs |
| --- | --- |
| Typical | More specific |
| Breathlessness<br>Orthopnoea<br>Paroxysmal nocturnal dyspnoea<br>Reduced exercise tolerance<br>Fatigue, tiredness, increased time to recover after exercise<br>Ankle swelling | Elevated jugular venous pressure<br>Hepatojugular reflux<br>Third heart sound (gallop rhythm)<br>Laterally displaced apical impulse |
| Less typical | Less specific |
| Nocturnal cough<br>Wheezing<br>Bloated feeling<br>Loss of appetite<br>Confusion (especially in the elderly)<br>Depression<br>Palpitations<br>Dizziness<br>Syncope<br>Bendopnea<sup>53</sup> | Weight gain (>2 kg/week)<br>Weight loss (in advanced HF)<br>Tissue wasting (cachexia)<br>Cardiac murmur<br>Peripheral oedema (ankle, sacral, scrotal)<br>Pulmonary crepitations<br>Reduced air entry and dullness to percussion at lung bases (pleural effusion)<br>Tachycardia<br>Irregular pulse<br>Tachypnoea<br>Cheyne Stokes respiration<br>Hepatomegaly<br>Ascites<br>Cold extremities<br>Oliguria<br>Narrow pulse pressure |

HF = heart failure.

### Appendix III CHS criteria for diagnosis of congestive heart failure^33^

**TABLE 4.** Definition of major cardiovascular events.

|  |  |
| --- | --- |
| Myocardial Infarction | Evolving Q-wave MI; *or* cardiac pain plus abnormal enzymes and either an evolving ST-T pattern or new left bundle branch block. |
| Angina Pectoris | Coronary artery bypass graft surgery; *or* $\geq 70\%$ obstruction of any coronary artery; *or* ST depression $> 1$ mm on exercise testing plus positive Rose questionnaire; *or* angina diagnosed by physician plus receiving medical treatment for angina (nitrates, beta-blockers or calcium-channel blockers). |
| Congestive Heart Failure | Cardiomegaly and pulmonary edema on chest X-ray; *or* dilated ventricle and wall-motion abnormalities by echocardiography or contrast ventriculography; *or* congestive failure diagnosed by physician, plus receiving medical treatment (diuretic plus either digitalis, vasodilator or angiotensin converting enzyme inhibitor). |
| Peripheral Arterial Disease | Ultrasonographically or angiographically demonstrated obstruction or ulcerated plaque; *or* absence of doppler pulse in any major vessel; *or* positive exercise test for claudication; *or* bypass surgery, angioplasty or thrombolysis for peripheral arterial disease; *or* exertional leg pain relieved by rest plus either claudication diagnosed by physician or ankle-arm systolic ratio of $\leq 0.8$. |
| Stroke | Abrupt onset of new neurologic deficit lasting at least 24 hours, with specific localizing findings confirmed by unequivocal physical examination or laboratory data and without evidence for underlying nonvascular cause. |
| Transient Ischemic Attack | Rapid onset of focal neurologic deficit lasting less than 24 hours, assessed to be due to ischemia, without evidence for underlying nonvascular cause. |

### Appendix IV Newcastle-Ottawa quality assessment scale for cohort studies^34^

#### NEWCASTLE - OTTAWA QUALITY ASSESSMENT SCALE COHORT STUDIES

<u>Note</u>: A study can be awarded a maximum of one star for each numbered item within the Selection and Outcome categories. A maximum of two stars can be given for Comparability

##### Selection

1. <u>Representativeness of the exposed cohort</u>
  a. truly representative of the average __________(describe) in the community ^**\***^
  b. somewhat representative of the average ________ in the community ^**\***^
  c. selected group of users eg nurses, volunteers
  d. no description of the derivation of the cohort
2. <u>Selection of the non exposed cohort</u>
  a. drawn from the same community as the exposed cohort^**\***^
  b. drawn from a different source
  c. no description of the derivation of the non exposed cohort
3. <u>Ascertainment of exposure</u>
  a. secure record (eg surgical records) ^**\***^
  b. structured interview^**\***^
  c. written self report
  d. no description
4. <u>Demonstration that outcome of interest was not present at start of study</u>
  a. yes^**\***^
  b. no

##### Comparability

1. <u>Comparability of cohorts on the basis of the design or analysis</u>
  a. study controls for ___________(select the most important factor) ^**\***^
  b. study controls for any additional factor (This criteria could be modified to indicate specific control for a second important factor.) ^**\***^

##### Outcome

1. <u>Assessment of outcome</u>
  a. independent blind assessment**\***
  b. record linkage**\***
  c. self report
  d. no description
2. <u>Was follow-up long enough for outcomes to occur</u>
  a. yes (select an adequate follow up period for outcome of interest)**\***
  b. no
3. <u>Adequacy of follow up of cohorts</u>
  a. complete follow up - all subjects accounted for**\***
  b. subjects lost to follow up unlikely to introduce bias - small number lost - > ____ % (select an adequate %) follow up, or description provided of those lost)**\***
  c. follow up rate < ____ % (select an adequate %) and no description of those lost
  d. no statement

### Appendix V Newcastle-Ottawa Scale (NOS) quality assessment of included case-control and cohort studies

| **Study**<br>**Authors** | **Newcastle-Ottawa**<br>**Scale** |  |  |  |
| --- | --- | --- | --- | --- |
|  | **Selection** | **Comparability** | **Outcome** | **Total** |
| Ho et al.,<br>2013 | 1-1-1-1 | 2 | 1-1-1 | 9 |
| Djoussé et al.,<br>2014 | 1-1-1-1 | 1 | 1-1-0 | 7 |
| Brouwers et<br>al., 2014 | 1-1-1-1 | 2 | 1-1-1 | 9 |
| Jagodzinski et<br>al., 2015 | 1-1-1-0 | 1 | 1-1-1 | 8 |
| AbouEzzedine<br>et al., 2016 | 1-1-1-1 | 1 | 1-1-1 | 8 |
| de Boer et al.,<br>2018 | 1-1-1-1 | 1 | 1-1-1 | 8 |
| Asleh et al.,<br>2019 | 1-1-1-0 | 1 | 1-1-1 | 7 |
| Bansal et al.,<br>2019 | 1-1-1-1 | 1 | 1-1-0 | 7 |
| Aguilar et al.,<br>2020 | 1-1-1-1 | 1 | 1-1-0 | 7 |

### Appendix VI Hazard ratios converted to 4^th^-to-bottom quartile of log-gal-3 and incident HF. Values not needing conversion are presented in their original form

| Study Authors | Converted Minimally Adjusted HR [95% CI] (Covariates) | Converted Further Adjusted HR [95% CI] | Covariates Used in Further Adjusted Model |
| --- | --- | --- | --- |
| Ho et al., 2013 | 2.31<br>[1.49, 3.59]<br>(Age, sex) | 1.69<br>[1.09-2.63] | Age, sex, SBP, anti-HTN treatment, BMI, diabetes mellitus, smoking, CAD, AF, BNP, and valvular heart disease. |
| Djoussé et al., 2014 | **Odds Ratio:**<br>1.60<br>[1.07-2.40]<br>(AF, BMI, diabetes) | **Odds Ratio:**<br>1.57<br>[1.03-2.39] | AF, BMI, diabetes, HTN, log(hsCRP), alcohol, exercise, and smoking. |
| Brouwers et al., 2014 | 1.52<br>[1.07-2.17]<br>(Age, sex) | 1.27<br>[0.87-1.86] | Age, sex, BMI, smoking, SBP, plasma glucose, total cholesterol. |
| Jagodzinski et al., 2015 | 1.46<br>[1.13-1.90]<br>(Region of Finland) | 1.22<br>[0.94-1.58] | Region of Finland, HDL/total cholesterol, SBP, anti-HTN treatment, smoking, diabetes, valvular heart disease, and eGFR. |
| AbouEzzedine et al., 2016 | 2.80<br>[2.02-3.90]<br>(Unadjusted) | 1.49<br>[0.97-2.29] | Age, sex, BMI, eGFR, HTN, SBP, CAD, and diabetes mellitus. |
| de Boer et al., 2018 | N/A | 1.19<br>[1.05-1.34] | Pooled and adjusted for age, sex, race/ethnicity, SBP, HTN, BMI, diabetes, smoking, LVH, left bundle branch block, and previous MI. |
| Asleh et al., 2019 | 4.25<br>[2.92-6.18]<br>(Age, sex) | 2.57<br>[1.74-3.8] | Age, sex, Charlson comorbidity index, maximum troponin T, and history of HF. |
| Bansal et al., 2019 | 1.49<br>[1.14-1.96]<br>(Age, sex, race, diabetes mellitus, CVD, smoking, 24-hr urinary protein, eGFR, SBP, BMI, LDL cholesterol, HDL cholesterol) | 1.33<br>[1.01-1.75] | Age, sex, race, diabetes mellitus, CVD, smoking, 24-hr urinary protein, eGFR, SBP, BMI, LDL cholesterol, HDL cholesterol, ACE inhibitors/blockers, diuretics, beta-blockers, phosphate, parathyroid hormone, FGF-23. |
| Aguilar et al., 2020 | 31.81<br>[9.94-101.86]<br>(Age, sex, race, SBP, anti-HTN treatment, smoking, diabetes mellitus, BMI, and heart rate) | 5.32<br>[1.43-19.85] | Age, sex, race, SBP, anti-HTN treatment, smoking, diabetes mellitus, BMI, heart rate, eGFR, log NT-proBNP, and log high sensitivity cardiac troponin T. |

**Abbreviations:** HR, hazard ratio; SBP, systolic blood pressure; HTN, hypertension; BMI, body-mass index; CAD, coronary artery disease; AF, atrial fibrillation; BNP, brain natriuretic peptide; CRP, C-reactive protein; HDL, high-density lipoprotein; eGFR, estimated glomerular filtration rate; LVH, left ventricular hypertrophy; MI, myocardial infarction; HF, heart failure; LDL, low-density lipoprotein; ACE, angiotensin-converting enzyme; FGF, fibroblast growth factor; NT-proBNP, N-terminal pro B-type natriuretic peptide.

## Notes

### Competing Interest Statement

The authors have declared no competing interest.

### Funding Statement

This study did not receive any funding

### Author Declarations

Since this was a meta-analysis of published data, all data was taken from previously published de-identified patient data (see Figure references 15, 18, 19, and 21-26 for original data which has already been published)

## REFERENCES

1. Heron, M. Deaths: Leading Causes for 2017. Natl. Vital Stat. Rep. Cent. Dis. Control Prev. Natl. Cent. Health Stat. Natl. Vital Stat. Syst. 68, 1–77 (2019).

2. Ahmad, F. B. & Anderson, R. N. The Leading Causes of Death in the US for 2020. JAMA 325, 1829–1830 (2021).

3. Inamdar, A. A. & Inamdar, A. C. Heart Failure: Diagnosis, Management and Utilization. J. Clin. Med. 5, (2016).

4. Baccouche, B. M. & Natterson-Horowitz, B. Giraffe Myocardial Hypertrophy as an Evolved Adaptation and Natural Animal Model of Resistance to Diastolic Heart Failure in Humans. (2019).

5. Natterson-Horowitz, B. et al. Did giraffe cardiovascular evolution solve the problem of heart failure with preserved ejection fraction? Evol. Med. Public Health 9, 248–255 (2021).

6. Baccouche, B. M. et al. The Burden of Heart Failure with Preserved Ejection Fraction in American Women is Growing: An Epidemiological Review. N. M. J. Sci. 55, (2021).

7. Savarese, G. & Lund, L. H. Global Public Health Burden of Heart Failure. Card. Fail. Rev. 3, 7–11 (2017).

8. Dong, R. et al. Galectin-3 as a novel biomarker for disease diagnosis and a target for therapy (Review). Int. J. Mol. Med. 41, 599–614 (2018).

9. Suthahar, N. et al. Galectin-3 Activation and Inhibition in Heart Failure and Cardiovascular Disease: An Update. Theranostics 8, 593–609 (2018).

10. Ziaeian, B. & Fonarow, G. C. Epidemiology and aetiology of heart failure. Nat. Rev. Cardiol. 13, 368–378 (2016).

11. Ilieşiu, A. M. & Hodorogea, A. S. Treatment of Heart Failure with Preserved Ejection Fraction. Adv. Exp. Med. Biol. 1067, 67–87 (2018).

12. Howard, B. E. et al. SWIFT-Review: a text-mining workbench for systematic review. Syst. Rev. 5, 87 (2016).

13. Baccouche, B. M. & Shivkumar, T. E. Using SWIFT-Review as a New and Robust Tool for Comprehensive Systematic Review. N. M. J. Sci. 54, (2020).

14. Moher, D., Liberati, A., Tetzlaff, J. & Altman, D. G. Preferred reporting items for systematic reviews and meta-analyses: the PRISMA statement. BMJ 339, b2535 (2009).

15. de Boer, R. A. et al. Association of Cardiovascular Biomarkers With Incident Heart Failure With Preserved and Reduced Ejection Fraction. JAMA Cardiol. 3, 215–224 (2018).

16. Kaptoge, S. Stata Programs. Cardiovascular Epidemiology Unit https://www.phpc.cam.ac.uk/ceu/erfc/programs/ (2005).

17. Stata | StataCorp LLC. https://www.stata.com/company/.

18. Asleh, R. et al. Galectin-3 Levels and Outcomes after Myocardial Infarction: A Community Study. J. Am. Coll. Cardiol. 73, 2286–2295 (2019).

19. Bansal, N. et al. Cardiac Biomarkers and Risk of Incident Heart Failure in Chronic Kidney Disease: The CRIC (Chronic Renal Insufficiency Cohort) Study. J. Am. Heart Assoc. Cardiovasc. Cerebrovasc. Dis. 8, (2019).

20. Chêne, G. & Thompson, S. G. Methods for summarizing the risk associations of quantitative variables in epidemiologic studies in a consistent form. Am. J. Epidemiol. 144, 610–621 (1996).

21. Ho, J. E. et al. Galectin-3, a Marker of Cardiac Fibrosis, Predicts Incident Heart Failure in the Community. J. Am. Coll. Cardiol. 60, 1249–1256 (2012).

22. Djoussé, L. et al. Plasma Galectin 3 and heart failure risk in the Physicians’ Health Study. Eur. J. Heart Fail. 16, 350–354 (2014).

23. Brouwers, F. P. et al. Clinical risk stratification optimizes value of biomarkers to predict new-onset heart failure in a community-based cohort. Circ. Heart Fail. 7, 723–731 (2014).

24. Jagodzinski, A. et al. Predictive value of galectin-3 for incident cardiovascular disease and heart failure in the population-based FINRISK 1997 cohort. Int. J. Cardiol. 192, 33–39 (2015).

25. AbouEzzeddine, O. F. et al. Biomarker-based risk prediction in the community. Eur. J. Heart Fail. 18, 1342–1350 (2016).

26. Aguilar, D. et al. Levels and Change in Galectin-3 and Association With Cardiovascular Events: The ARIC Study. J. Am. Heart Assoc. Cardiovasc. Cerebrovasc. Dis. 9, (2020).

27. International Classification of Diseases, Ninth Revision. https://www.cdc.gov/nchs/icd/icd9.htm (2015).

28. Mähönen, M. et al. The validity of heart failure diagnoses obtained from administrative registers. Eur. J. Prev. Cardiol. 20, 254–259 (2013).

29. Alhajj, M. & Farhana, A. Enzyme Linked Immunosorbent Assay. in StatPearls (StatPearls Publishing, 2021).

30. Cinquanta, L., Fontana, D. E. & Bizzaro, N. Chemiluminescent immunoassay technology: what does it change in autoantibody detection? Auto-Immun. Highlights 8, (2017).

31. McKee, P. A., Castelli, W. P., McNamara, P. M. & Kannel, W. B. The Natural History of Congestive Heart Failure: The Framingham Study. N. Engl. J. Med. (1971) doi:http://dx.doi.org/10.1056/NEJM197112232852601.

32. Ponikowski, P. et al. 2016 ESC Guidelines for the diagnosis and treatment of acute and chronic heart failure: The Task Force for the diagnosis and treatment of acute and chronic heart failure of the European Society of Cardiology (ESC)Developed with the special contribution of the Heart Failure Association (HFA) of the ESC. Eur. Heart J. 37, 2129–2200 (2016).

33. Fried, L. P. et al. The Cardiovascular Health Study: design and rationale. Ann. Epidemiol. 1, 263–276 (1991).

34. Wells, G. et al. Ottawa Hospital Research Institute. The Newcastle-Ottawa Scale (NOS) for assessing the quality of nonrandomised studies in meta-analyses http://www.ohri.ca/programs/clinical_epidemiology/oxford.asp (2015).

